# Adaptive data-driven age and patch mixing in contact networks with recurrent mobility

**DOI:** 10.1101/2021.09.29.21264319

**Authors:** Jesse Knight, Huiting Ma, Amir Ghasemi, Mackenzie Hamilton, Kevin Brown, Sharmistha Mishra

## Abstract

Infectious disease transmission models often stratify populations by age and geographic patches. Contact patterns between age groups and patches are key parameters in such models. Arenas et al. (2020) develop an approach to simulate contact patterns associated with recurrent mobility between patches, such as due to work, school, and other regular travel. Using their approach, mixing between patches is greater than mobility data alone would suggest, because individuals from patches A and B can form a contact if they meet in patch C. We build upon their approach to address three potential gaps that remain. First, our approach includes a distribution of contacts by age that is responsive to underlying age distribution of the mixing pool. Second, different age distributions by contact type are also maintained in our approach, such that changes to the numbers of different types of contacts are appropriately reflected in changes to the overall age mixing patterns. Finally, we introduce and distinguish between two mixing pools associated with each patch, with possible implications for the overall connectivity of the population: the home pool, in which contacts can only be formed with other individuals residing in the same patch; and the travel pool, in which contacts can be formed with some residents of, and any other visitors to the patch. We describe in detail the steps required to implement our approach, and present results of an example application.

**Graphical Abstract:** 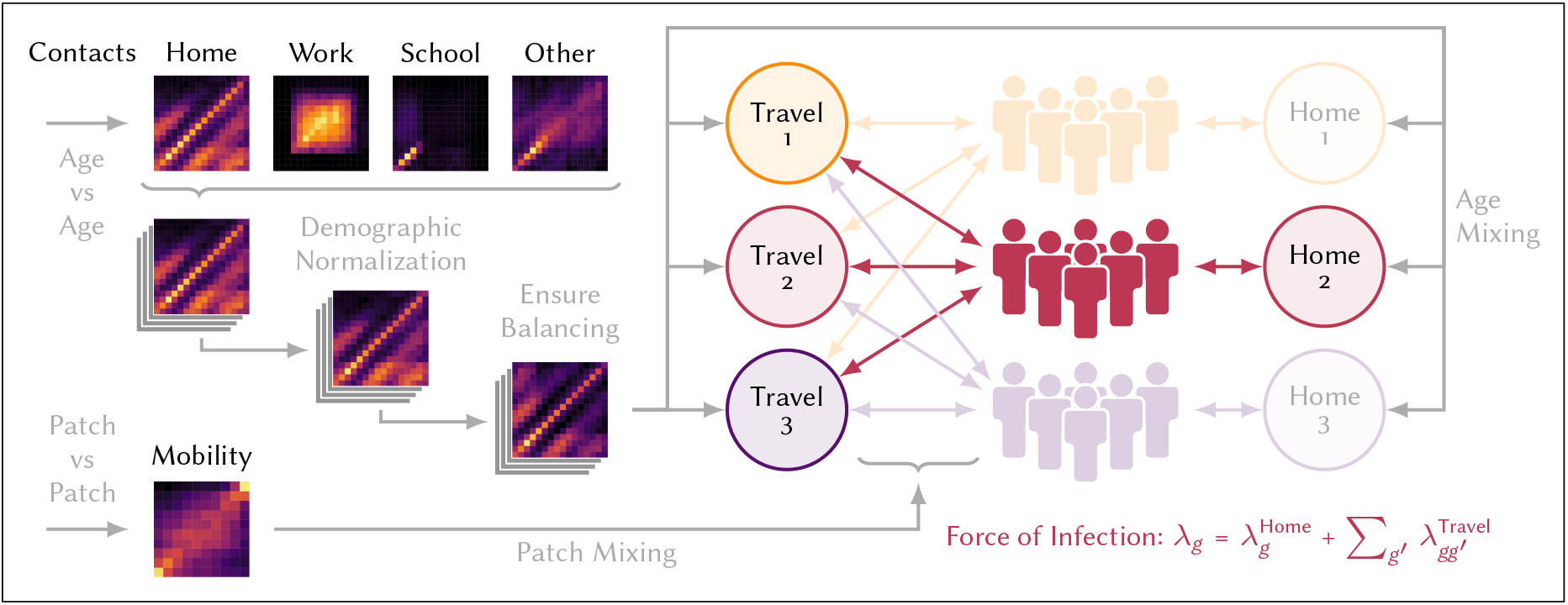

## 1 Introduction

Contact patterns are key determinants of epidemic dynamics because they define who can be infected, by whom, and how quickly [2]. Arenas et al. [1] develop a patch-based model of sars-cov-2 transmission applied to Spain, in which the modelled population is stratified by geographic patches and three age groups. Following foundational work by Balcan and Vespignani [3] and Sattenspiel and Dietz [4], the model incorporates data on short, recurrent mobility patterns to determine contact rates between individuals in different patches and age groups. We build upon this contact model to incorporate improved age mixing patterns, which are stratified by different contact types and are responsive to the age distributions of mixing populations, as proposed by Arregui et al. [5]. We also explore some practical considerations in parameterizing such models.

## 2 Method

Consider a population stratified by *N*_*g*_ patches and *N*_*a*_ age groups.^1^ Let *P*_*ga*_ be the number of people in patch *g* and age group *a*. Let *y* denote *N*_*y*_ different types of contacts (e.g. household, workplace, etc.). Let *B*_*gg*′_ be the proportion of population *P*_*g*_ who travel to *g*^′^ each day, or the “mobility matrix”.^2^

### 2.1 Original Approach

Arenas et al. [1] model the force of infection (incidence per susceptible) experienced by population *P*_*ga*_ as:

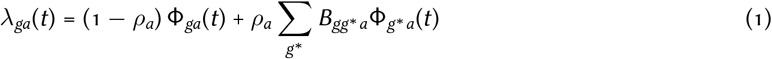

where: Φ_*ga*_(*t*) is the probability of acquiring infection while in patch *g*; and *ρ*_*a*_ ∈ [0, 1] is an age-specific overall mobility factor. Thus, *λ*_*ga*_(*t*) is the sum of infection probabilities from the residence patch *g*, and from visited patches *g*^∗^ ≠ *g*. The probability Φ_*g*∗ *a*_(*t*) is modelled using the chained binomial for multiple exposures [6]:

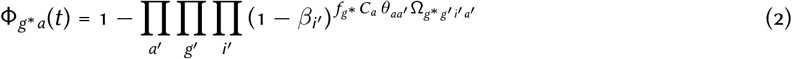

where: *β*_*i*_ is the per-contact transmission probability associated with infectious state *i*; *f*_*g*∗_ is a density factor associated with patch *g*^∗^; *C*_*a*_ is the expected number of contacts made per person per day in age group *a*; *θ*_*aa*′_ is the age distribution of those contacts, derived from [7] for Spain, such that ∑_*a*′_ *θ*_*aa*′_ = 1; and Ω_*g*∗ *g*′*i*′*a*′_ is the proportion of individuals present in patch *g*^∗^ who reside in patch *g*^′^ and who are in infectious state *i*^′^, for each age group *a*^′^. This proportion Ω_*g*∗ *g*′*i*′*a*′_ is defined as:

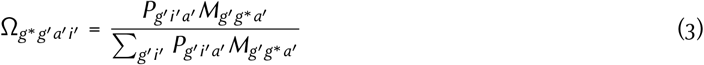

where *M*_*gg*′*a*_ is a convenience simplification of the mobility matrix:

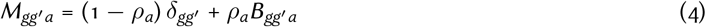

This model of infection captures important mixing patterns related to recurrent mobility that are relevant to epidemic modelling on relatively small spatial and time scales. However, the model could be improved by separating different contact types throughout the force of infection equation, and by allowing age mixing patterns to respond to local demographic and intervention conditions. Three specific issues with the original approach are as follows:

1. **Contact balancing:** The contact balancing principle states that the total number of contacts formed by group *a* with group *a*^′^ should equal the number formed by group *a*^′^ with group *a* [5]:

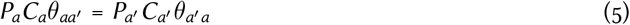 For a model with non-random age mixing and random (proportionate) mixing by patches, Eq. (5) could be satisfied by a single fixed age mixing matrix *θ*_*aa*′_, i.e. for the population overall. However, in the context of patch-based mixing reflecting recurrent mobility, Eq. (5) should be satisfied in each mixing context (patch). Specifically, if different patches have different age distributions, or different rates of per-person contact formation due to household size, employment, etc., then it would not be possible to satisfy Eq. (5) with a single fixed age mixing matrix *θ*_*aa*′_. The implications of violating Eq. (5) depend on relative differences in demographics and/or contact rates by patch and/or age group. For example, if a given patch skews younger than average in age, and most contacts are formed with other members of the same patch, then fixed average *θ*_*aa*′_ would underestimate the number of younger contacts among residents of this patch, and overestimate the number of older contacts.
2. **Age mixing by contact type:** A related issue is that the expected contact rates by age group *C*_*a*_ reflect the summation of different types of contacts, and so the fixed age mixing matrix *θ*_*aa*′_ is applied to all contact types. As such, changes to the numbers of each type of contact are not paired with changes the overall mixing patterns. As illustrated by the polymod study [2], age mixing patterns vary by contact type, such as highly age-assortative mixing in schools. Thus, differential reductions in each contact type would affect overall age mixing patterns. For example, if reductions in school-related contacts due to school closures were not reflected in *θ*_*aa*′_, then the relative contribution of children to overall transmission could be overestimated during the period of school closures.
3. **Modelling contact & mobility reductions:** The term (1 − *ρ*_*a*_) Φ_*ga*_(*t*) in Eq. (1) represents transmission to non-mobile individuals in patch *g*. The associated definitions in Eqs. (2–4) consider transmission to these non-mobile individuals from visitors to patch *g*. Such definitions therefore imply that non-mobile individuals still form contacts with visitors to their residence patch. However, it may be useful to model some or all non-mobile individuals as only forming contacts with other individuals from their residence patch. That is, scenarios may exist wherein a fraction of the population only has household contacts, as could be the case with public health measures such as lockdowns. As illustrated in Figure A.1, the original approach may overestimate inter-patch connectivity during periods of reduced mobility (via lockdowns) versus an approach in which some or all non-mobile populations are limited to contacts with others from their residence patch and not with visitors. Thus, the original approach [1] could underestimate the impact of confinement lockdown strategies on inter-patch transmission reduction.

We therefore develop a refinement of the original approach, with the aim of addressing the above three issues.

### 2.2 Proposed Approach

In the proposed approach, the contributions of different contact types to the force of infection are added to the binomial function for multiple exposures:

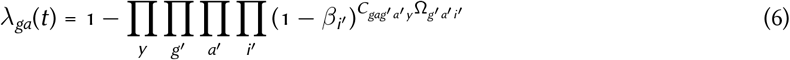

where: *C*_*gag*′*a*′*y*_ is the expected number of type *y* contacts formed per person per day among individuals in population *P*_*ga*_ with those in population *P*_*g*′*a*′_ ; and Ω_*g*′*a*′*i*′_ is the proportion of individuals in residing in patch *g*^′^ and age group *a*^′^ who are in infectious state *i*^′^:

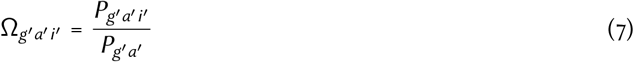

For each type of contact, *C*_*gag*′*a*′*y*_ is defined to reflect both age-related and mobility-related mixing factors, as described in the following subsections. To support these descriptions, we will refer to Figures from an example application, although the details of the application and the Figures are given in § 3. Collecting the full network of contacts in the matrix *C*_*gag*′*a*′*y*_ provides a representation that is easy to interpret, and allows us to compute various properties, like the margins in *a, a*^′^ or *g, g*^′^, and whether contact balancing is satisfied per Eq. (5). Additionally, separating contact types allows the incorporation of different probabilities of transmission per contact type *β*_*i*′*y*_, if desired.

#### 2.2.1 Age Mixing

Prem et al. [8] project contact patterns by 5-year age groups from the polymod study [2] onto 177 countries, considering various demographic data. These contact matrices represent *C*_*aa*′*y*_ : the expected number of type *y* contacts formed per day among individuals in age group *a* with those in age group *a*^′^. Four types of contact are considered: “home”, “work”, “school”, and “others”^3^ (Figure 4a). We aim to incorporate these contact numbers and patterns into *C*_*gag*′*a*′*y*_.

The first challenge is that the contact matrices *C*_*aa*′*y*_ are inherently weighted by the underlying population age distribution—the proportion of expected contacts with age group *a*^′^ is proportional to the size of age group *a*^′^. To overcome this challenge and apply these patterns to new population age structures, Arregui et al. [5] suggest to divide by the population age distribution to obtain an “unweighted” matrix 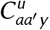 (Figure 4b):^4^

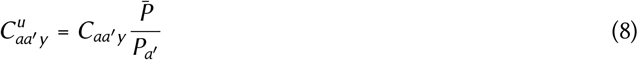

where 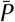 is the mean of *P*_*a*′_.

The next challenge is that 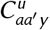 may not satisfy the contact balancing principle, Eq. (5), due to sampling and/or reporting error in the polymod survey. To ensure that the overall mixing matrix *C*_*gag*′*a*′*y*_ will satisfy the balancing principle, the input age mixing matrix 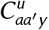 must satisfy the principle. A simple solution is to average 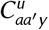 with its transpose to obtain the “balanced” matrix 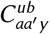 (Figure 4c):

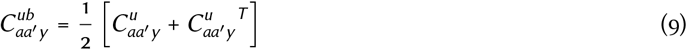

This operation may change the margin *C*_*ay*_, representing the total type *y* contacts formed by individuals in age group *a*. However, such changes are reasonable if understood as a correction for sampling bias.^5^

A final challenge in applying the contact matrices from [8] is that the 5-year age groups may not align with the age groups of interest. Overcoming this challenge is not theoretically required to obtain *C*_*gag*′*a*′*y*_, but we describe a solution here in case it is useful for modelling applications. We begin by upsampling the contact matrix from 5-year age groups *a*_5_ to 1-year age groups *a*_1_ using bilinear interpolation, based on the midpoints of each age group, and scaled by a factor of 1*/*5. To avoid edge effects associated with many interpolation implementations, we first pad the matrix by replicating the edges diagonally. If the desired age groups extend beyond the maximum age group of 80 available in [8], diagonal padding can also be used to approximate the trends in the additional age groups. Then, given the age groups of interest *a*_∗_ (which may have irregular widths), we aggregate 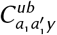 to obtain 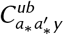 using matrix multiplication with indicator matrix *A*:

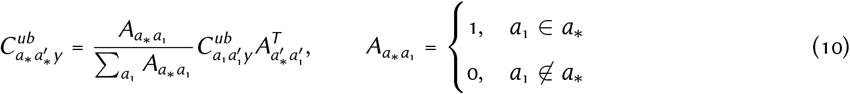

The right-hand *A*^*T*^ term *sums* the total number of contacts formed with the 1-year “other” age groups 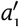 corresponding to 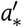. The left-hand *A* term *averages* the total number of contacts formed from the 1-year “self” age groups *a*_1_ corresponding to *a*_∗_. The average weights each 1-year age group *a*_1_ equally, although other weights could be incorporated through the nonzero values of *A*. Another interpretation of the normalization sum is the widths of the age groups *a*_∗_.

The resulting matrix 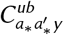 represents the expected contacts among age groups *a*_∗_ when mixing with a population having equal proportion in all age groups 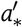 (regardless of their width). Thus, 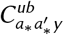 can later be multiplied by the population age distribution of interest—reversing Equation (8)—to obtain the expected number of contacts when mixing with that population. This approach then addresses issues 1 and 2 described in § 2.1.

#### 2.2.2 Mobility-Related Mixing

In conceptualizing mobility-related mixing, we define two types of contexts in which contacts can be formed:

- **Home pools:** where contacts are formed exclusively with other residents of the same patch (e.g. for household contacts)
- **Travel pools:** where contacts are formed with individuals from any patch who are present in the pool (e.g. for work contacts)

We model one home pool and one travel pool associated with each patch, as illustrated in Figure 1.

**Figure 1:**
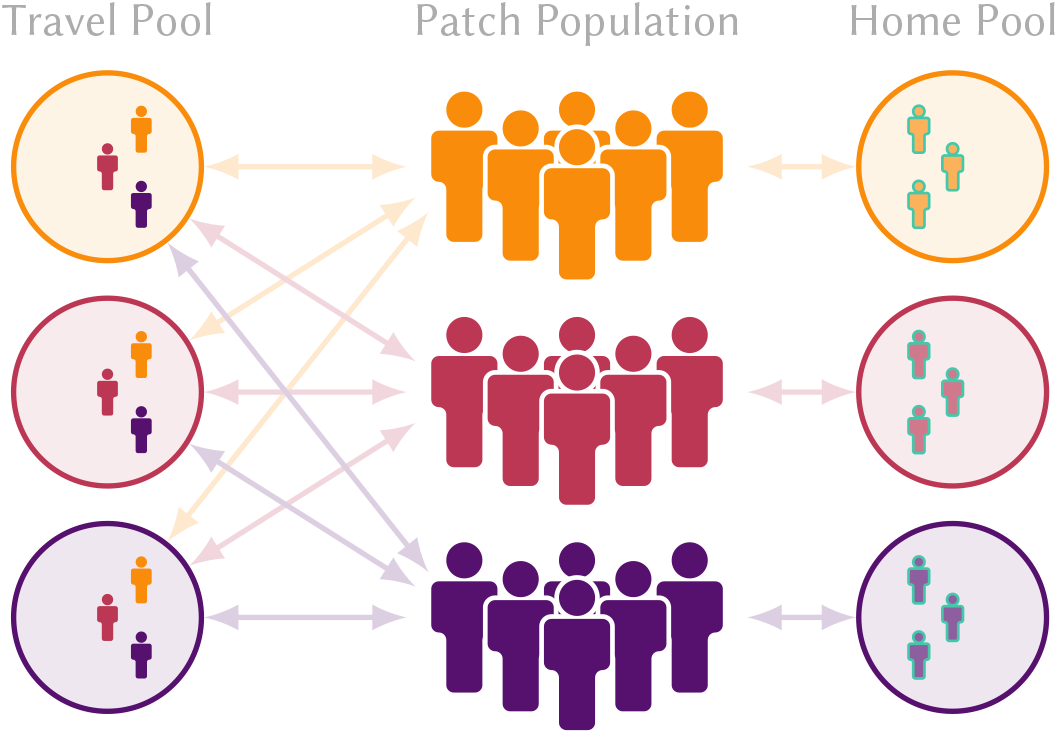
Toy example of “home” vs “travel” mixing pools for a network with 3 patches and 50% individuals mobile. Contacts in the home pool are formed exclusively with other members of the residence patch, whereas contacts in the travel pool may be formed with any visitors to the patch Non-mobile populations are indicated with faded colour and green outline

In this conceptualization, only contacts associated with travel pools are influenced by the population mobility matrix *B*_*gg*_′, representing the expected proportions of individuals from patch *g* who visit patch *g*^′^ per day. For contacts associated with home pools, this matrix is functionally replaced with an identity matrix *δ*_*gg*_′. It is not necessary to assume that all contacts of any particular type are formed in only one type of pool. Rather, we introduce a parameter *h*_*y*_ ∈ [0, 1] representing the proportion of type *y* contacts that are formed in the home pool, and the remainder (1 − *h*_*y*_) are formed with travel pools. For example, we could have *h*_*y*_ = 1 for household contacts, *h*_*y*_ = 0 for work contacts, and *h*_*y*_ = 0.5 for school contacts. Thus, the expected contacts formed by individuals in patch *g* are distributed across three situations:

1. **Mobile Away:** individual travelled from patch *g* to patch *g*^′^ and formed contacts within travel pool *g*^′^ Proportion of contacts: (1 − *h*_*y*_)*B*_*gg*_′ (g ≠ *g*′)
2. **Mobile at Home:** individual formed contacts within their local travel pool *g* Proportion of contacts: (1 − *h*_*y*_)*B*_*gg*_′ _(*g*=*g*′)_
3. **Non-Mobile at Home:** individual formed contacts within their local home pool *g* Proportion of contacts: *h*_*y*_ *δ*_*gg*_′

The idea of “home pools” is new versus [1], and allows us to address issue 3 by introducing situation 3. Thus in [1], all mixing was implicitly modelled using “travel pools”, and individuals described as “non-mobile” reflected situation 2.

In the context of reduced mobility, we do not assume that rows of *B*_*gg*_′ sum to 1. The “missing” proportion 1 − Σ_*g*_′ *B*_*gg*_′ is then taken to represent non-mobile individuals, who do not form any mobility-related contacts (situations 1 and 2) that day. In § A.3 we discuss some details about generating a mobility matrix *B*_*gg*_′ with these properties, based on mobile phone data.

To calculate *C*_*gag*_′_*a*_′_*y*_ using these assumptions, we begin by considering the travel pool in patch *g*^∗^. The effective number of individuals from population *P*_*ga*_ who are present in the pool is given by:^6^

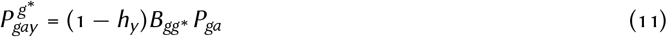

There is no distinction between situations 1 and 2 in Eq. (11), as both are already reflected in the off-diagonal and diagonal elements of *B*_*gg*_′, respectively. If we assume that mixing by residence patch *g* within the pool is random, we need only consider age mixing within the pool. Under completely random mixing and with 1 contact per person, the total number of contacts formed between 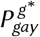 and 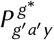 is given by the outer product:

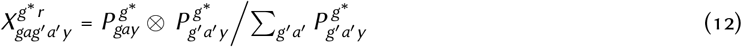

where the first term represents the absolute population size of “self”, and the second term represents proportions of their contacts among “other” strata. Then, the numbers and patterns of contacts by age can be applied via multiplication:

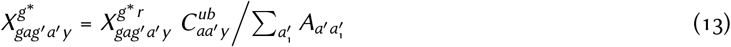

since 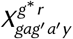 is proportional to the population age distribution of “others”, and will therefore act to reverse Eq. (8) as planned. The term 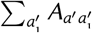 is from Eq. (10), representing the widths of the age groups *a*^′^. It is and necessary to divide by the widths of age groups *a*^′^ since both 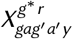 are 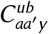 proportional to these widths, but the proportionality should only be singular overall. We could have applied this normalization to 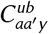 in Eq. (10) in the same way as for *a*, but this would make 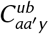 harder to interpret, as it would no longer represent the expected numbers of contacts for each age group.

Mixing within home pools (situation 3) can be modelled similar to mixing within travel pools, with one small modification: replacing (1 − *h*_*y*_)*B*_*gg*_′ with *h*_*y*_ *δ*_*gg*_′. Following through Eqs. (11–12), we obtain 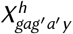, representing the total contacts formed within home pools. Then, the total type *y* contacts formed between populations *P*_*ga*_ and *P*_*g*_′_*a*_′ across all relevant mixing pools is given by the sum:

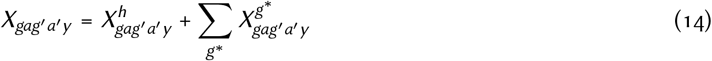

It may be tempting to simplify the model for home pool contacts by updating the mobility matrix *B*_*gg*_′ similar to Eq. (4) from [1], with *h*_*y*_ = (1 − *ρ*_*a*_). However, such an approach does not produce the same result as Eq. (14), and indeed underpins issue 3 described in § 2.1 regarding mixing of non-mobile individuals with mobile visitors to their patch. On the other hand, if the interpretation of “non-mobile” is intended to allow mixing with visitors, then *B*_*gg*_′ can still be adjusted per Eq. (4) to simulate this behaviour. Another implication of our approach is that non-mobile individuals will not form mobility-related contacts. Thus, if Σ_*g*_′ *B*_*gg*_′ is reduced, the total contacts formed by residents of patch *g* would be reduced proportionately, and changes to mixing patterns by age and patch reflected automatically.

Finally, the number of type *y* contacts formed *per person* in population *P*_*ga*_ with population *P*_*g*_′_*a*_′ can be obtained by dividing *X*_*gag*_′_*a*_′_*y*_ by the population size:

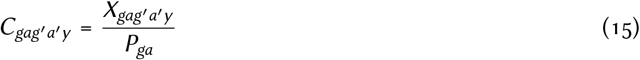

## 3 Example

We applied the proposed methodology for generating a mixing matrix *C*_*gag*_′_*a*_′_*y*_, which reflects patterns of age mixing, recurrent mobility between patches, and different contact types, to the population (14 million) of Ontario, Canada, in the context of covid-19 transmission modelling. Ten patches were defined based on groupings of the 513 forward sortation areas (FSAs)^7^ in Ontario. The FSA groupings reflect deciles of cumulative covid-19 cases, excluding cases among residents of long-term are homes, between 15 January 2020 and 28 March 2021 [9]. Thus each patch represents approximately 10% of the Ontario population (37–68 FSAs), but not contiguous geographic regions. Such definitions were used to support allocation and prioritization of covid-19 vaccines to “hot spot” neighbourhoods in Ontario [10, 11]. Figure 2 illustrates the locations of the FSAs and their decile rank, which is synonymous with their patch index. Figure A.10 plots the daily incidence of covid-19 cases per patch, and Figure A.11 plots the age distributions of each patch. Age groups were then defined to reflect historical and hypothetical covid-19 vaccine eligibility in Ontario:

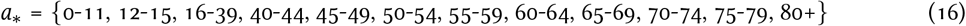

**Figure 2:**
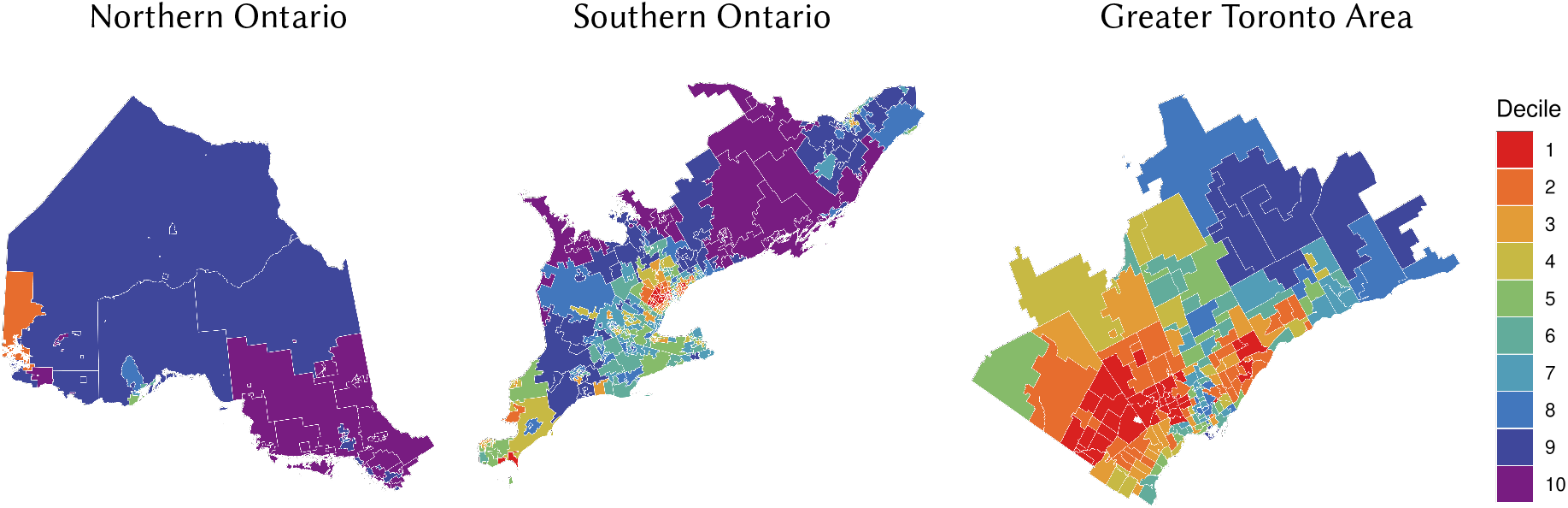
Ontario forward sorting areas (FSAs, *N* = 513), stratified by decile rank in cumulative covid-19 cases between 15 Jan 2020–28 Mar 2021; decile rank was used to group FSAs into 10 patches for transmission modelling.

### 3.1 Data

Ontario population sizes by age and FSA *P*_*ga*_ were obtained from the 2016 Canadian Census via Statistics Canada^8^ and aggregated from 1-year age groups (*a*_1_) into 5-year (*a*_5_) and target (*a*_∗_) age groups as needed. We obtained the final output contact matrices *C*_*aa*_′_*y*_ for Canada from [8], for each of the “home”, “work”, “school”, and “others” contact types, as well as the population size of each 5-year age group used in [8].^9^ We assumed that residence patch did not influence the numbers of contacts formed per person, only with whom those contacts are formed, although such a belief could be incorporated in the model, perhaps in Eq. (13).

The mobility matrix *B*_*gg*_′ between patches was derived using private data on geolocation service usage among a sample of approximately 2% of mobile devices in Ontario [12] during January–December 2020. Appendix A.3 details the specific methods and assumptions used; to summarize: Each devices was assigned an approximate home location (152.9 m × 152.4 m) based on the most common location during overnight hours for each calendar month. This location was then used to determine the home FSA (*n*). The proportion of time spent outside the home location each day, stratified by inside vs outside the home FSA, was also used to estimate the relative proportions of intra-vs inter-FSA mobility. Finally, the total numbers of visits to other FSAs (*n*^′^) by all devices were used to estimate the conditional probability of travelling from FSA *n* to FSA *n*^′^, given that an individual will travel outside the home FSA *n*.

The contribution of each FSA to overall mobility of the patch/decile (group of FSAs) was then aggregated as:

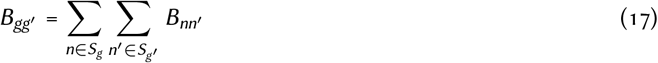

where *S*_*g*_ is the set of FSAs (*n*) corresponding to patch/decile *g*. Mobility matrices were estimated for each month in the available dataset (Jan–Dec 2020). A reference period reflecting pre-pandemic conditions was defined as Jan–Feb 2020; unless otherwise specified, all subsequent results use the average mobility patterns during that period (Figure 3). We did not model any differences in mobility by age group, although such differences could be included in the model by adding a relative rate in Eq. (11).

**Figure 3:**
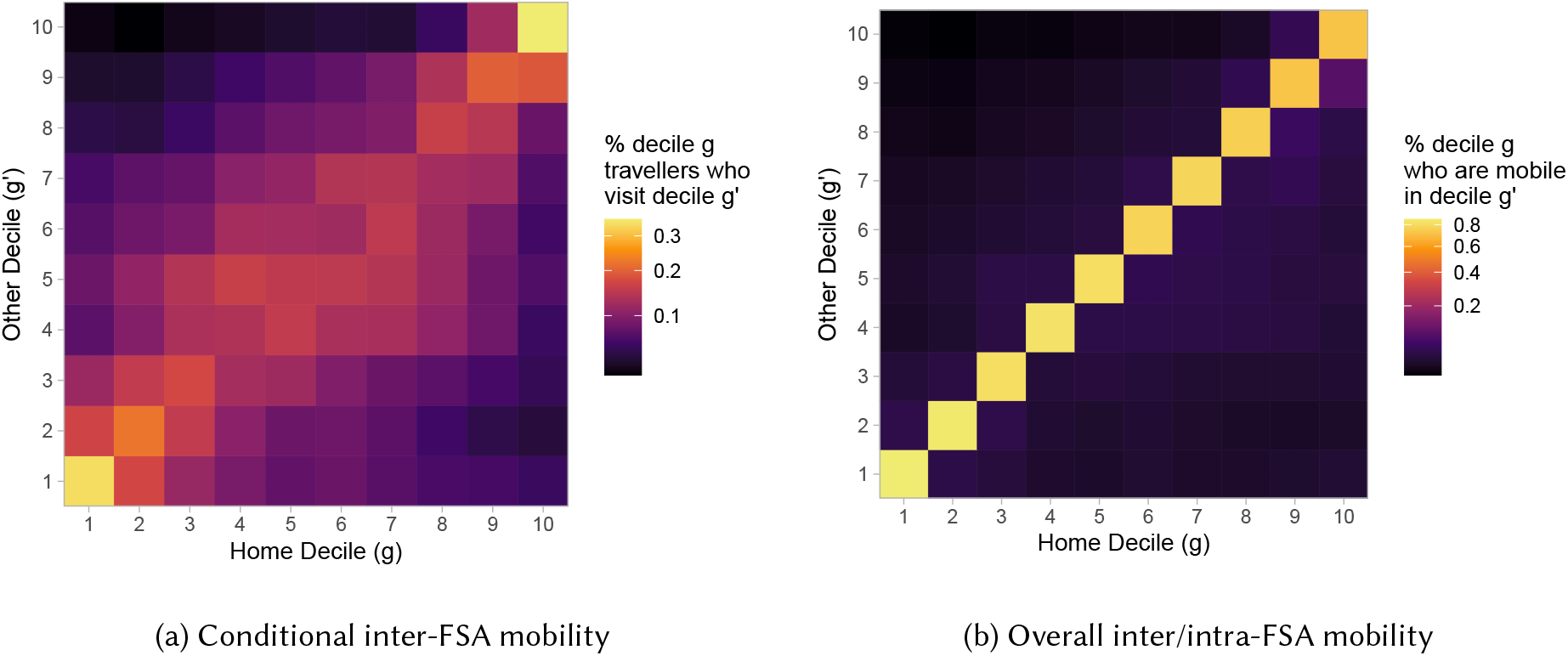
Mobility matrix *B*_*gg*_′, representing the expected proportion of individuals in decile (patch) *g* who are mobile in decile *g*^′^ per day Derived from mobile device geolocation data; deciles represent groupings of Ontario forward sortation areas (FSAs) by cumulative covid-19 cases between 15 Jan 2020–28 Mar 2021; colour scale is square-root transformed to improve perception of smaller values; reference period: Jan–Feb 2020.

Finally, we specified the proportions of each contact type assumed to be formed with the home pool:

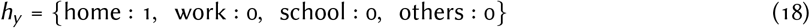

The parameters *P*_*ga*_, *C*_*aa*_′_*y*_, *B*_*gg*_′, and *h*_*y*_ represent the necessary inputs to our approach for calculating *C*_*gag*_′_*a*_′_*y*_. The following § 3.2 walks step-wise through the approach and presents all major intermediate results.

### 3.2 Results

Figure 4 illustrates the contact matrices *C*_*aa*_′_*y*_ from Prem et al. [8], before and after the steps of unweighting by population age distributions, Eq. (8), and ensuring contact balancing, Eq. (9). Figure 5 illustrates the differences in contact matrices between each step. These differences can be explained as follows. The Canadian age distribution used by Prem et al. [8] (Figure A.11, black dashed line), is below the mean for the youngest and oldest age groups; thus inverting the weighting by this age distribution increases the contacts expected with these age groups (Figure 5a). By contrast, Figure 5b is purely symmetric (and opposite about the central diagonal), reflecting differences from the symmetric mean matrix.

**Figure 4:**
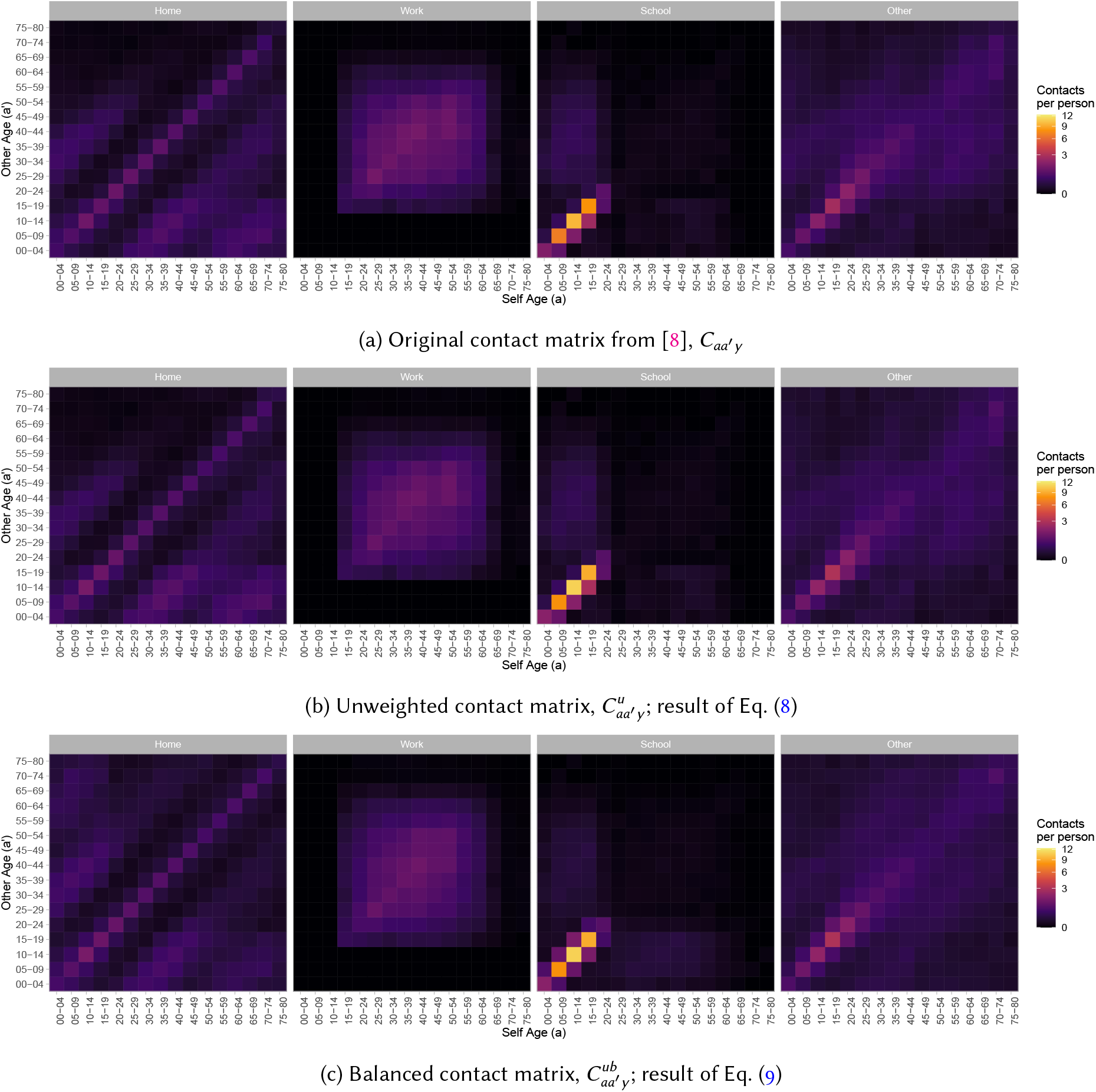
Intermediate results in obtaining unweighted and balanced age contact matrices 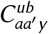 (expected number of type *y* contacts per person per day in each age group *a*, with those other age groups *a*^′^) from population-weighted matrices *C*_*aa*_^′^_*y*_ from [8] which may not satisfy contact balancing Contact matrices for Canada, derived from [8]; colour scales are square-root transformed to improve perception of smaller values.

**Figure 5:**
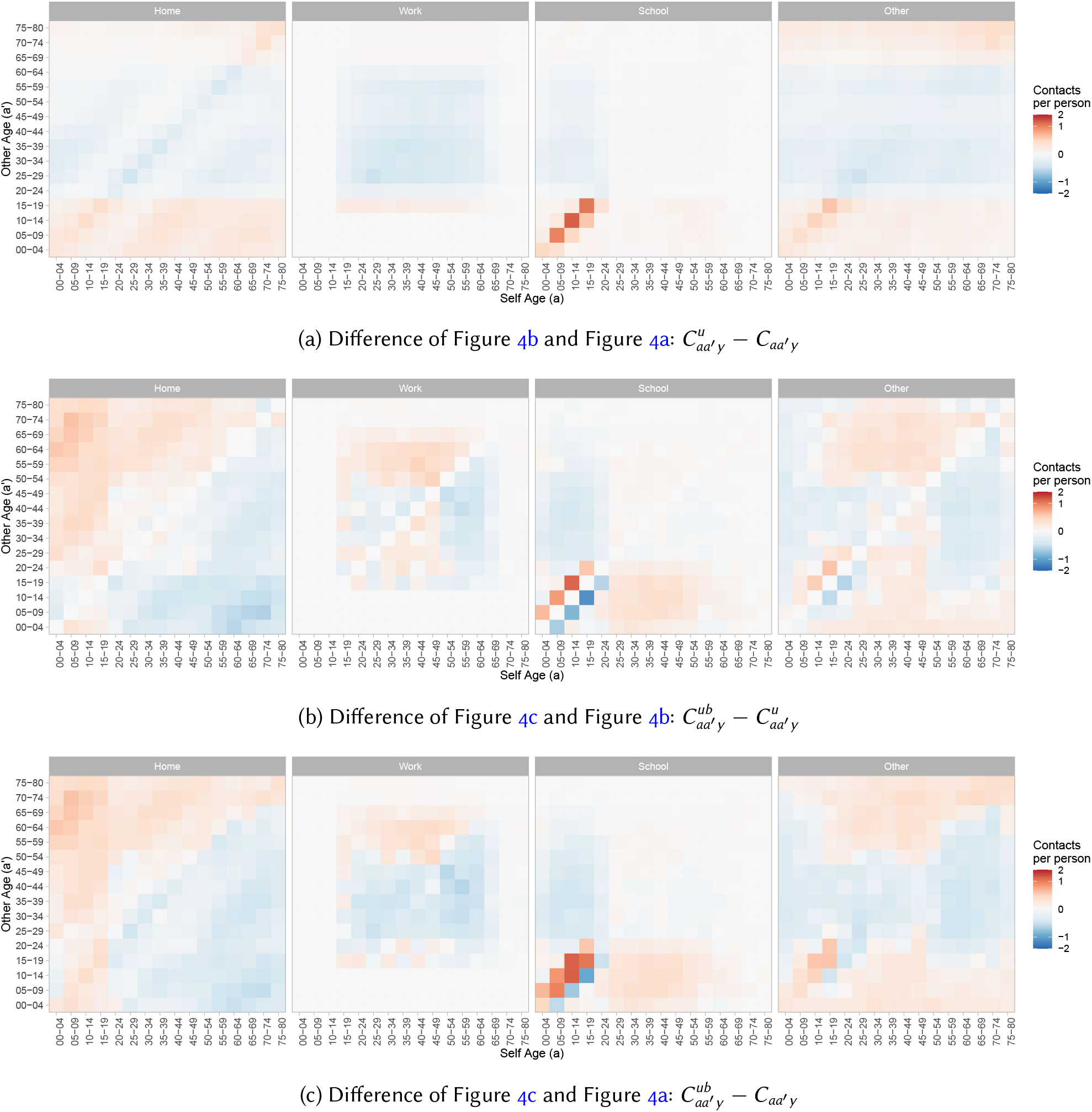
Differences between intermediate results shown in Figure 4 Contact matrices for Canada, derived from [8]; colour scales are square-root transformed to improve perception of smaller values.

Figure 6 illustrates the unweighted and balanced contact matrices 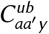 before and after bilinear interpolation and aggregation to the target age groups of interest, Eq. (10). The final matrices 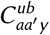 include dominant horizontal streaks corresponding to larger age groups. These streaks are expected, as more contacts are expected to form with larger “other” age groups. Vertical streaks do not appear, as each column represents the expected contacts for each person in the “self” age group, not the total contacts formed by that age group.^10^

**Figure 6:**
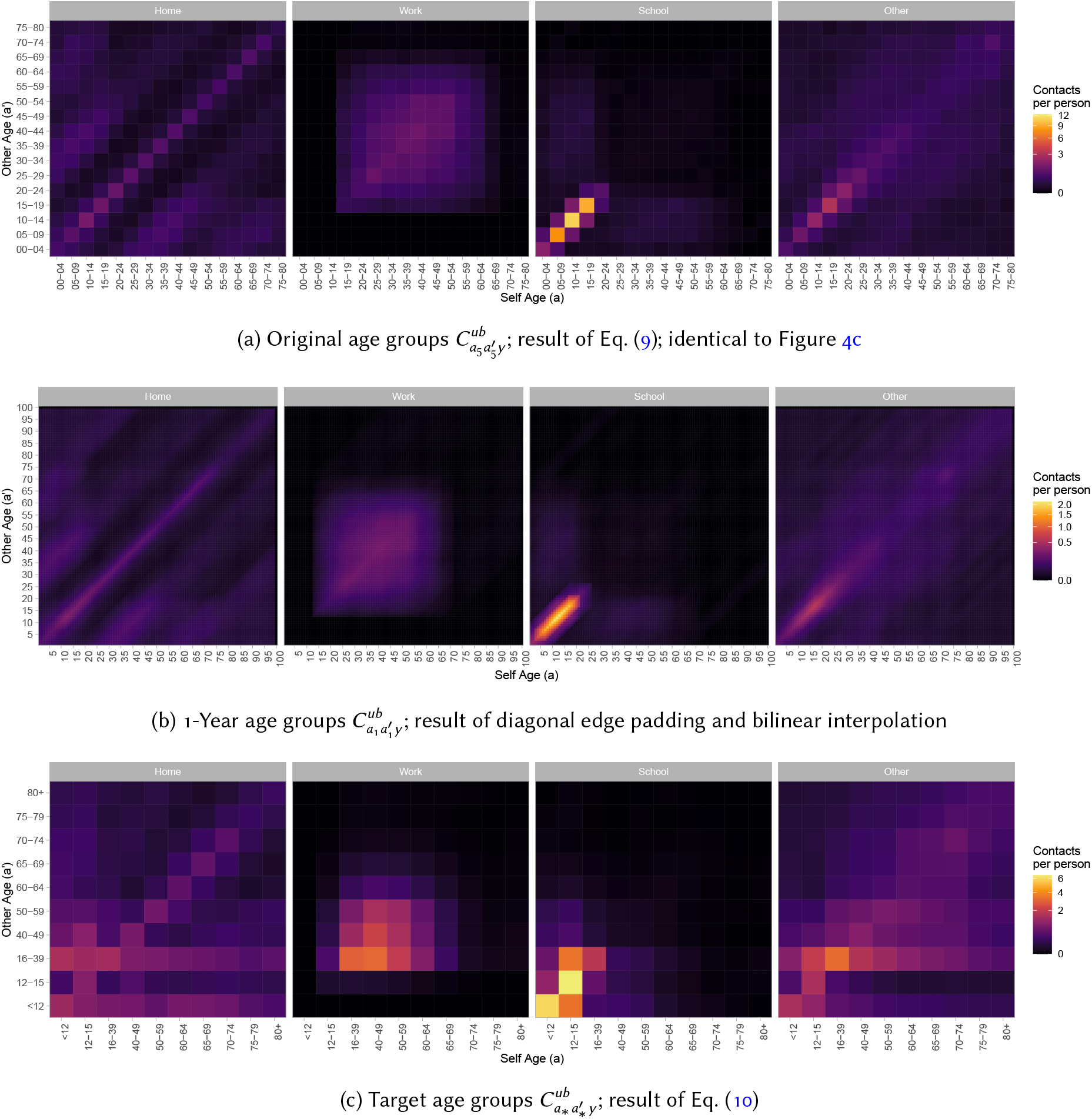
Intermediate results in obtaining age-restratified contact matrices 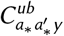 (expected number of type *y* contacts per person per day in each age group *a*_∗_, with those from age groups 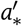) from matrices 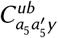 with year age stratifications *a*_5_ Contact matrices for Canada, derived from [8]; colour scales are square-root transformed to improve perception of smaller values; the horizontal streaks in (c) corresponding to age groups 0–11 and 16–39 are expected, as more contacts will be formed with larger age groups.

Figure 7 plots the total expected contacts per person per day, *C*_*ay*_ = Σ_*a*_′ *C*_*aa*_′_*y*_, before and after each of the above steps, from before Eq. (8) through after Eq. (10). Overall, patterns remained roughly consistent across transformations, although some details among the large 16–39 age group are lost due to substantial averaging.

**Figure 7:**
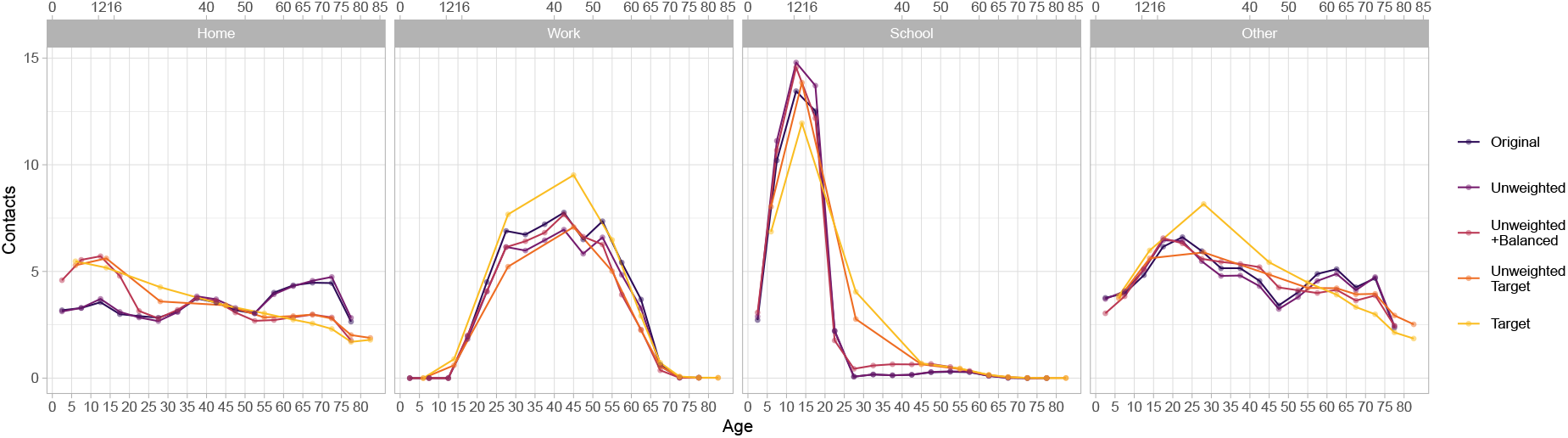
Total contacts per person per day *C*_*ay*_ = Σ_a_^′^ *C*_*aa*_^′^_*y*_ for each intermediate step in obtaining 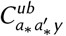 stratified by contact type. Contact matrices for Canada, derived from [8]; modelled contacts for each age group are plotted at the midpoint of the age group; the cut points for the original age groups *a*_5_ from [8] and the target age groups *a*_∗_ in our application are indicated on the bottom and top x-axes, respectively.

Finally, Figure 8 illustrates the margins (sum over “other” strata and population-weighted average over “self” strata) of the complete mixing matrices *C*_*gag*_′_*a*_′_*y*_, in terms of age groups *a* & *a*^′^ (Figure 8a), and patches/deciles *g* & *g*^′^ (Figure 8b). Such margins are computed as follows:

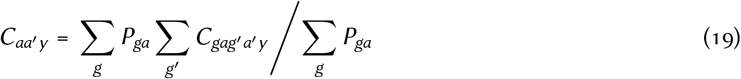

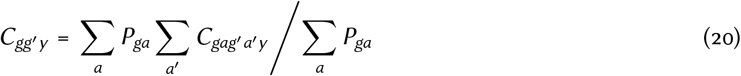

**Figure 8:**
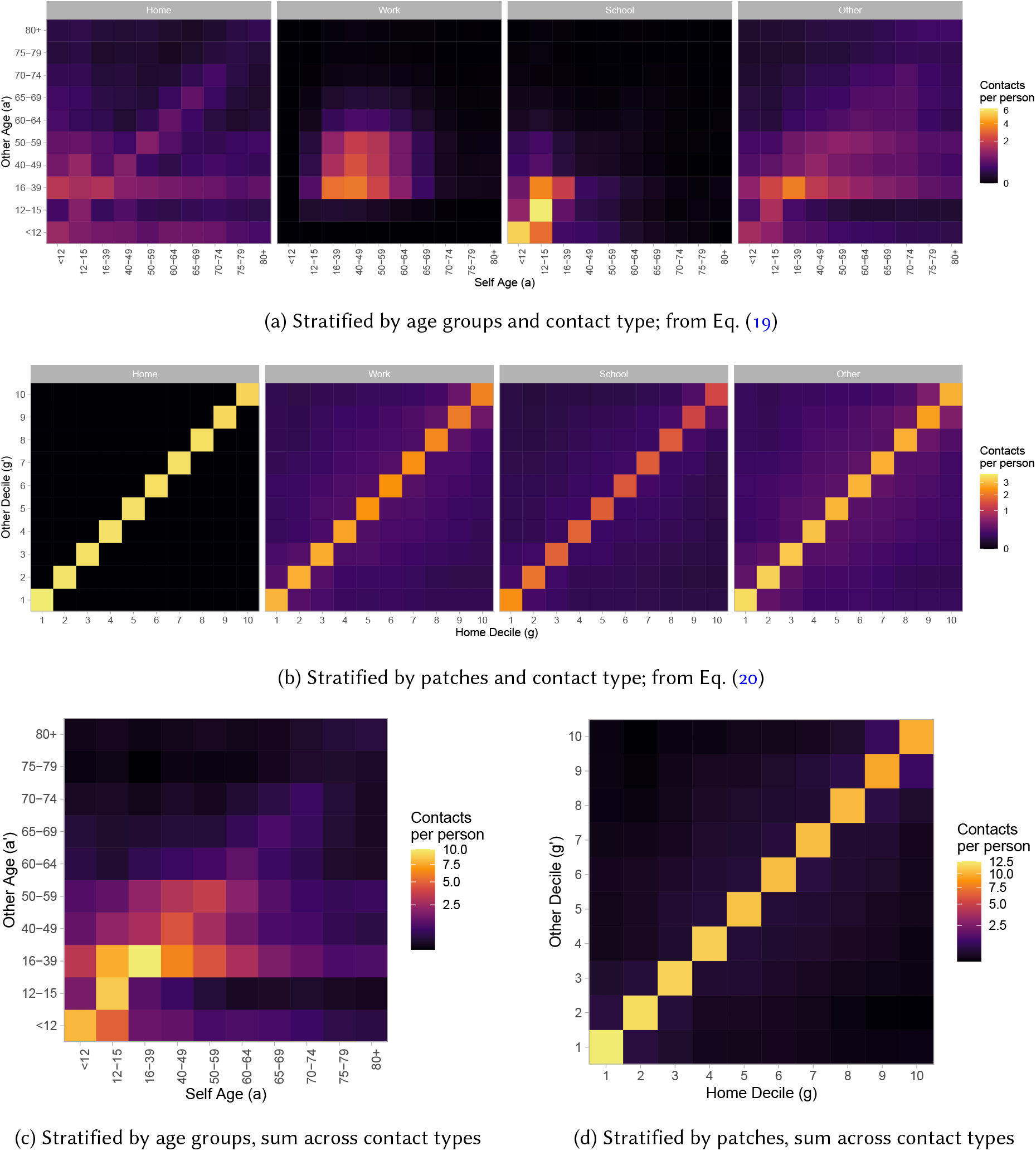
Expected contacts per person per day, stratified by age, decile (patch), and contact type, computed as the margins of the overall contact matrices *C*_*gag*_^′^_*a*_^′^_*y*_ Contact matrices for Canada, derived from [8]; colour scales are square-root transformed to improve perception of smaller values.

The equivalent matrices for total number of contacts per person of all types (*C*_*aa*_′ and *C*_*gg*_′) are also given in Figures 8c and 8d, respectively. The marginal matrices *C*_*aa*_′_*y*_ are identical to the input age mixing matrices from Eq. (10), which could be used as an implementation check. Since *h*_*y*_ = 1 for “home” contacts, *C*_*gg*_′_*y*_ is an identity matrix. The equivalent matrices for “work”, “school”, and “others” contact types also feature a strong diagonal, due to a strong diagonal in the source mobility matrix *B*_*gg*_′ (individuals who are mobile within their home FSA). However, the off-diagonal elements are less clustered towards the central diagonal versus the mobility matrix *B*_*gg*_′ (Figure 3). Mobile individuals from patches *g* and *g*^′^ may form contacts not only when either travels to the others’ patch, but also when they both travel to a third patch. Thus, the degree of mixing between patches using this approach is greater than the mobility matrix alone would suggest, though less than if completely random mixing was assumed.

## 4 Conclusion

Arenas et al. [1] develop an approach to modelling contact patterns associated with recurrent mobility, which is relevant to dynamic models of infectious disease transmission, such as sars-cov-2. The original approach simulates contact patterns between age groups and geographic patches connected by recurrently mobile individuals, and considers changes to mixing between patches due to reduced mobility among some individuals. In this paper, we proposed approaches to improve upon the approach to: ensure contact balancing between age groups; model changes to age contact patterns in response to reduced mobility; and allow complete isolation of non-mobile individuals from mobility-related contacts.

The first key change in the proposed approach draws on [5] to combine preferential patterns of age mixing with the age distribution of the mixing population, such that the actual number of contacts formed reflects both elements. This change is incorporated into each separate mixing “pool” where contacts are formed, and ensures that the number of contacts simulated from age group *a* to age group *a*^′^ will equal those from age group *a*^′^ to age group *a*. This revised approach can also ensure contacts balance if population sizes change over time, such as in the case of large differential mortality by age group or patch; or if the numbers of contacts formed per-person differ by patch, such as if individuals in some patches have higher numbers of work contacts.

The second key change in the proposed approach is to maintain separate mixing patterns for each type of contact, only aggregating the contribution of different contact types to overall transmission within the force of infection equation. With this change, the age mixing patterns associated with any contact type are not influenced by changes to the numbers or mixing patterns of any other contact type. This change also supports differential probability of transmission by contact type.

The final key change in the proposed approach is to introduce two separate mixing pools where contacts can form. Within “home” pools, contacts can only be formed with other residents of the same patch. Within “travel” pools, contacts cab be formed with other residents of the same patch who are mobile within their residence patch, or with any mobile visitors to the patch. Home pools therefore allow true isolation of some individuals from mobility-related contacts, with implications for overall network connectivity.

In developing and applying the proposed approach to an example context, we present the methodological details and results of each intermediate step, so that they may be reproduced or built upon in future work.

## Data Availability

All analysis code and selected outputs for the study are available online at the link below. Where possible, input data have also been made publicly available in the same repo, although this was not possible for several variables / stratifications.

https://github.com/mishra-lab/age-patch-mobility-mixing

## Acknowledgements

We thank: Kristy Yiu (Unity Health Toronto) for research coordination support; Gary Moloney (Unity Health Toronto) for support with geographic data processing and helpful discussions; and Linwei Wang (Unity Health Toronto) for helpful discussions.

Reported covid-19 cases were obtained from the Public Health Ontario (PHO) integrated Public Health Information System (iPHIS), via the Ontario covid-19 Modelling Consensus Table, and with approval from the University of Toronto Health Sciences REB (protocol No. 39253).

## Contributions

JK conceived of and developed the approach, reviewed the literature, conducted the mixing and mobility analyses, and wrote the code and manuscript. AG conducted and led data analyses and developed the mobility metrics. HM provided critical input into the assumptions and bias adjustments with the mobility data. MH, KB, and SM contributed critical insights to the assumpitons, data, approach, and interpretation. All authors were involved in intepretation and manuscript revisions.

## Footnotes

1 We use different notation than Arenas et al. [1]; a comparison is given in Table A.1.

2 Arenas et al. [1] consider different mobility patterns by age: *B*_*gg*′*a*_. For simplicity, we consider *B*_*gg*′_ unstratified by age, but age stratification could be added to our approach.

3 The “others” contact type in [8] is itself derived from the combination of “leisure”, “transport”, and “other” contact types from [2], while the “home”, “work”, and “school” types are the same between the studies.

4 The matrix 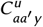 could also be interpreted as the expected contact matrix for a population with a rectangular demographic pyramid [5].

5 A perfect survey in a closed population would produce a contact matrix 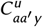 that is already balanced.

6 If residents of different patches might have relatively different numbers of contacts, a scaling factor could be applied here.

7 Each FSA is the first 3 characters of the postal code.

8 https://www150.statcan.gc.ca/n1/en/catalogue/98-400-X2016008

9 The rows for Canada in `contacts home.rdata`, `contacts work.rdata`, `contacts school.rdata`, and `contacts others.rdata` from `/generate synthetic matrices/output/syntheticcontactmatrices2020/` and `poptotal.rdata` from `/generate synthetic matrices/input/pop/` within https://github.com/kieshaprem/synthetic-contact-matrices/tree/6e0eebc.

10 The corresponding matrix of absolute contact *X*_*aa*_′_*y*_ = *C*_*aa*_′_*y*_ *P*_*a*_′ will be symmetric, and thus will include symmetric horizontal and vertical streaks.

