## Appendix for "Adaptive data-driven age and patch mixing in contact networks with recurrent mobility"

**Resource:** https://github.com/mishra-lab/age-patch-mobility-mixing

**Date:** Sept 29, 2021

### A Supporting Materials

#### A.1 Notation

Table A.1 summarizes our notation, and any corresponding parameters from [1].

Table A.1: Notation

| Symbol in this | Symbol in [1] | Definition |
|---|---|---|
| $g$ | $i$ | home patch |
| $g'$ | $j$ | other patch |
| $g^*$ | — | visited patch |
| $a$ | $g$ | self age group |
| $a'$ | $h$ | other age group |
| $y$ | — | contact type |
| $n$ | — | home FSA |
| $n'$ | — | visited FSA |
| $i$ | $m$ | infection state |
| $P$ | $N$ | population size |
| $B$ | $R$ | mobility matrix |
| — | $M$ | convenience mobility matrix |
| $\lambda$ | $\Pi$ | force of infection |
| $\rho$ | $p$ | mobility factor |
| $h$ | — | proportion of contacts formed at home |
| $C$ | $k$ | contacts per person |
| $X$ | — | total absolute contacts in the model |
| $\theta$ | $C$ | age distribution of contacts |
| $\Psi$ | — | proportion of time away from home |
| $\phi$ | — | proportion of away time in home FSA |

#### A.2 Interpretation of "Non-mobile"

As described in § 2.1, Arenas et al. [1] model the degree of mobility of age group $a$ using a parameter $\rho_a$ (our notation), incorporated into a convenience matrix $M_{gg'}$ per Eq. (4) / (A.1), repeated here for reference:

$$M_{gg'a} = (1 - \rho_a)\, \delta_{gg'} + \rho_a B_{gg'a} \tag{A.1}$$

The implicit assumption of this approach is that non-mobile individuals may form contacts with visitors to the former's residence patch (situation 2 in § 2.2.2): an assumption which may not be desired depending on the research question or intervention effect. The proposed approach to modelling mixing can avoid this assumption if needed through the use of "home pools" (situation 3). Figure A.1 explores the potential influence of compulsory mixing of non-mobile individuals with mobile visitors on network connectivity, as measured by the expected proportion of contacts formed with other patches, in a toy example. The example has 3 patches, each having equal population size and random mobility ($B_{gg'} = 1/N_{g'}$). Age is not considered.

By simulating mixing between non-mobile individuals and mobile visitors, as described in issue 3 (Figures A.1b & A.1d, situation 2) the proportions of contact made with other patches increases for all patches, as compared to the proposed approach (Figures A.1a & A.1c, situation 3). The difference is largest in the context of differential mobility by patch (Figures A.1d vs A.1c). Thus, the original approach could underestimate the transmission reduction impacts of reduced mobility, especially when some patches can reduce mobility-related mixing while others cannot.

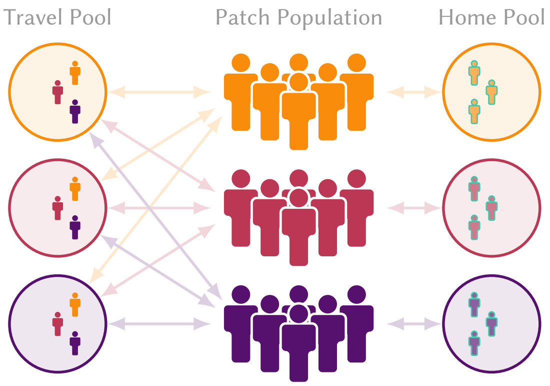


(a) Non-mobile = stay at home, all patches 50% mobile

Contacts formed with other patches:
33% if non-mobile have equal contact rates vs mobile
44% if non-mobile have 1/2 contact rates vs mobile

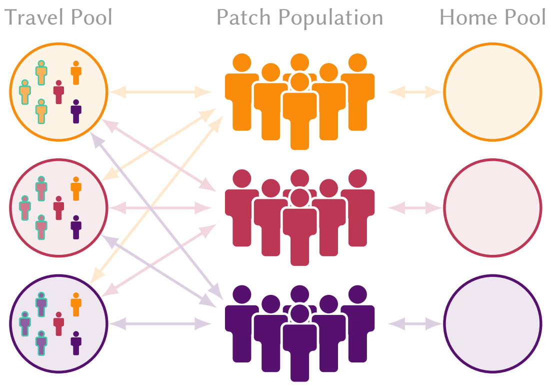


(b) Non-mobile = stay in patch, all patches 50% mobile

Contacts formed with other patches:
50% if non-mobile have equal contact rates vs mobile
59% if non-mobile have 1/2 contact rates vs mobile

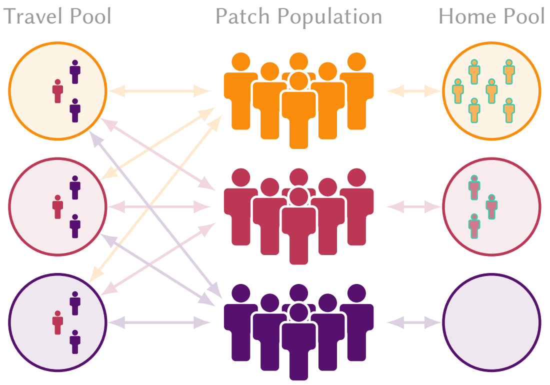


(c) Non-mobile = stay at home, patches 0, 50, 100 % mobile

Contacts formed with other patches:
0, 33, 33 % if non-mobile have equal contact rates vs mobile
0, 44, 33 % if non-mobile have 1/2 contact rates vs mobile

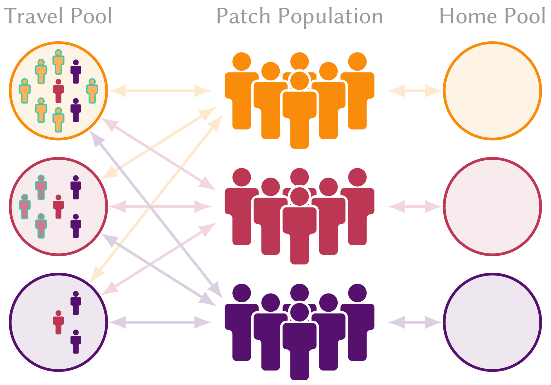


(d) Non-mobile = stay in patch, patches 0, 50, 100 % mobile

Contacts formed with other patches:
33, 48, 59 % if non-mobile have equal contact rates vs mobile
50, 58, 52 % if non-mobile have 1/2 contact rates vs mobile

Figure A.1: Mixing implications for two assumptions about the behaviour of non-mobile populations (issue 3): "stay at home" (a,c, reflecting situation 3): no contacts with other patches (proposed here); vs "stay within patch" (b,d, reflecting situation 2): no travel, but may form contacts with travellers from other patches (as in [1]).

Non-mobile populations are indicated with faded colour and green outline; patch-specific values ordered from top (yellow) to bottom (purple). Additional assumptions: large population size; equal population sizes and contact rates by patch; random mobility (equal probability of visiting any patch if mobile).

#### A.3 Deriving the Mobility Matrix: Example

Here we describe the methods used to obtain the mobility matrix $B_{gg'}$ for our applied example in Ontario Canada (details in § 3). The mobility matrix represents the expected proportions of individuals residing in decile (patch) $g$ who travel to decile $g'$ each day.

##### A.3.1 Data

The data are from a private anonymized database representing approximately 2% of mobile devices in Ontario.[11] The raw data represent logs of: unique device ID, timestamp, and geolocation (latitude / longitude, with accuracies of meters to tens of meters). A log entry ("ping") is generated when an app on the device requests the current geolocation from the service provider. The geolocation is compared to: a) boundary files representing all 513 Ontario FSAs[12] to determine which FSA the ping is attributed to; and b) Geohash-7 tiles (152.9 m × 152.4 m) to help identify the approximate home location. The following definitions were then used in determining device mobility.

- **Home dwelling:** for each unique device, the Geohash-6 tile with the greatest total evening time (8:00 pm–5:00 am) each calendar month. Devices for which it was not possible to determine the home dwelling were excluded from all further analysis.
- **Home FSA:** for each device, the FSA containing the midpoint of the home dwelling Geohash-6 tile.
- **Visited FSA:** FSAs that a device travelled to within a 24-hour period, as defined by at least 2 consecutive pings spanning at least 2 hours within the FSA. By this definition, it was possible for a device to visit multiple FSAs, or no FSA during a given day. Repeated visits to the same FSA by the same device on the same calendar date are treated as a single visit.
- **Time away from home:** for each device, the proportion of total time spent away from the home dwelling. Only devices with at least 5 pings spanning at least 4 hours were included.
- **Time away from home within home FSA:** for each device, the proportion of total time spent away from the home dwelling but still within the home FSA.

These definitions were applied to each calendar month $t$ from Jan 2020–Dec 2020 (12 months) to compute: the mean number of devices with home FSA $n$ per day, denoted $H_{nt}$ or "observed devices"; the mean number of devices with home FSA $n$ that visited FSA $n'$ per day, denoted $V_{nn't}$, or "device visits"; the mean proportion of time each device spent outside the home per day for each FSA, denoted $\Psi_{nt}$; and the mean proportion of time each device spent outside the home but within the home FSA per day for each FSA, denoted $\Psi^h_{nt}$

##### A.3.2 Mobility Matrix

The inter-FSA mobility matrix $B_{nn't}$ ($n \neq n'$) could be defined as the expected proportion of observed devices that travelled to each FSA per day ($V_{nn't}/H_{nt}$). However, the following analysis of the data suggested that

[11] https://www.veraset.com

[12] https://www150.statcan.gc.ca/n1/en/catalogue/92-179-X

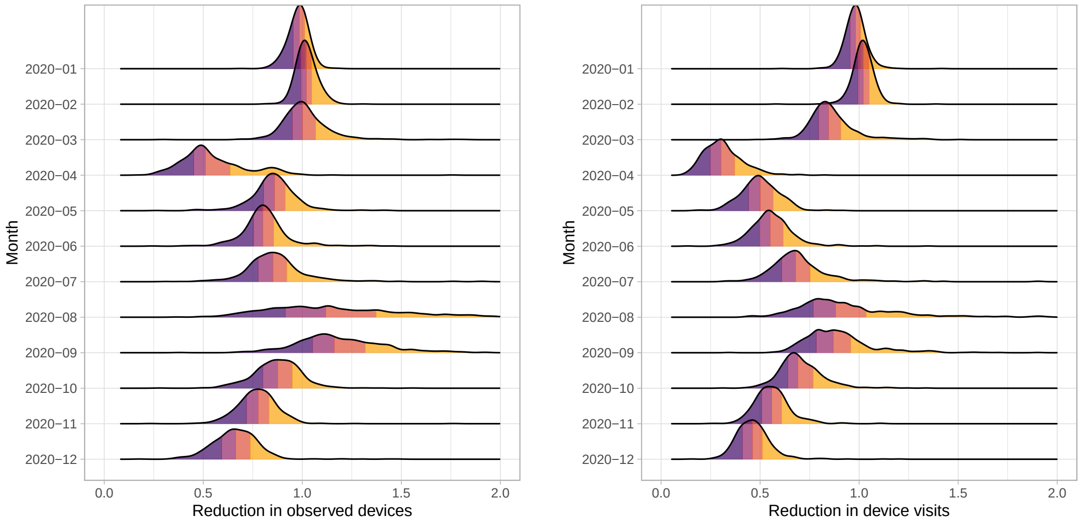


(a) Observed devices at home

(b) Total device visits

Figure A.2: Ratios of (a) observed devices at home and (b) total device visits during each month versus during the reference period suggesting that the proportion of observed devices travelling each month alone is not reflective of true mobility or else (a) would be 1

Data from approximately 2% of mobile devices in Ontario, Canada; distributions show the density of ratio values across all 513 FSAs; coloured segments indicate the 4 quantiles of each distributions; reference period: Jan–Feb 2020.

such an approach may bias estimates of mobility. A reference period $t_0$ was defined as Jan–Feb 2020, to reflect pre-pandemic conditions. The expected values of $H_{nt}$ and $V_{nt} = \sum_{n'} V_{nn't}$ for each FSA $n$ during this period were then computed and compared to the expected values during each subsequent month. Figure A.2 plots the distribution of ratios $H_{nt}/H_{nt_0}$ (a) and $V_{nt}/V_{nt_0}$ (b) across all 513 FSAs, for each month. These ratios illustrate that both $V_{nt}$ and $H_{nt}$ were influenced (reduced) by pandemic restrictions, and thus $V_{nn't}/H_{nt}$ would overestimate mobility. The trend in $H_{nt}$ might be because apps accessing geolocation services are only opened after the user intends to travel, such as map-related apps. As such, we term this bias "mobility-intention bias".

To address the mobility-intention bias, we separated the probability of an individual being mobile overall $\rho_{nt}$ from the conditional probability of travelling to FSA $n'$ given that the individual is going to travel outside their home FSA $B^c_{nn't}$. We defined this conditional probability of visiting FSA $n'$, given that the individual is travelling outside the home FSA as:

$$B^c_{nn't} = \frac{V_{nn't}}{\sum_{n'} V_{nn't}}, \qquad n \neq n' \tag{A.2}$$

which no longer considers the total number of devices observed $H_{nt}$. We then aggregated $B^c_{nn't}$ to obtain $B^c_{gg't}$ as noted in § 3.1:

$$B^c_{gg't} = \sum_{n \in S_g} \sum_{n' \in S'_g} B^c_{nn't} \tag{A.3}$$

where $S_g$ is the set of FSAs ($n$) corresponding to decile $g$. Figure A.3 illustrates the conditional probabilities over the available months.

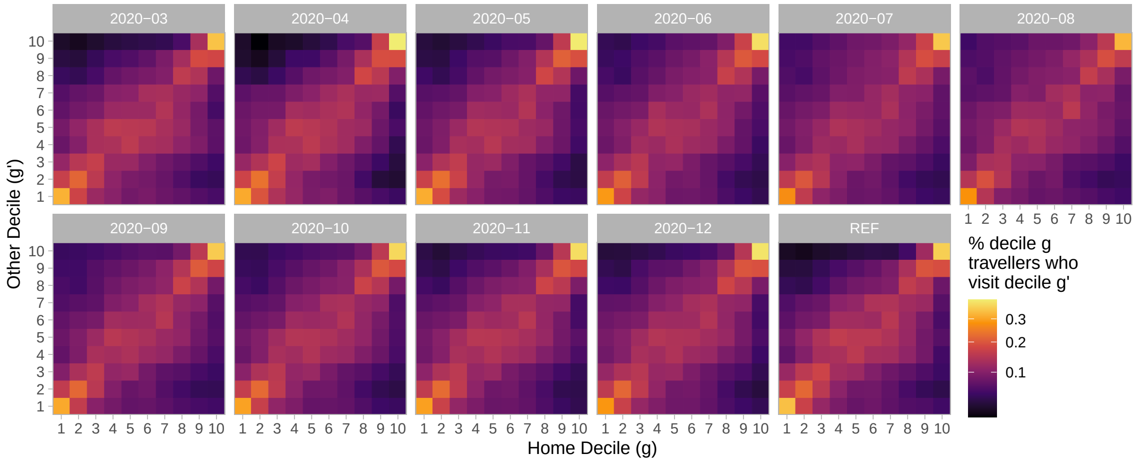


Figure A.3: Conditional probabilities $B^c_{gg't}$ of travelling from decile $g$ to decile $g'$ for Mar–Dec 2020, plus REF ($t_0$) as the average of Jan–Feb 2020

Derived from mobile device geolocation data; deciles represent groupings of Ontario forward sortation areas (FSAs) by cumulative COVID-19 cases between 15 Jan 2020–28 Mar 2021; colour scale is square-root transformed to improve perception of smaller values

Next, we defined the overall probability of being mobile outside the home for individuals from FSA $n$ during month $t$ as:

$$\rho_{nt} = \frac{\Psi_{nt}}{\Psi_{nt_0}} \tag{A.4}$$

reflecting an assumption that 100% of individuals are mobile outside the home during the reference period $t_0$, and thus $\rho_{nt}$ represents a relative reduction from this baseline. This definition of $\rho_{nt}$ aims to avoid the mobility-intention bias because it is not directly affected by the numbers of observed devices each month ($V_{nn't}$ and/or $H_{nt}$), but rather the behaviour (relative change in time away from home) associated with those devices. Even if observed devices are more mobile than unobserved devices, we think the reduction in time away from home relative to $t_0$ among observed devices/individuals would be reasonably representative of the reduction among unobserved devices/individuals.

Similar to $\rho_{nt}$, we then defined the proportion of time away from home spent within the home FSA as:

$$\phi_{nt} = \frac{\Psi^h_{nt}}{\Psi_{nt}} \tag{A.5}$$

Figure A.4 illustrates the distribution of $\rho_{nt}$ (b) and $\phi_{nt}$ (b) across different FSAs, stratified by month and decile. We observe that the lower incidence deciles had the lowest relative reduction in mobility $\rho_{nt}$ (b), and sometimes had greater mobility during the pandemic as compared to the reference period. We also observe that the majority of time spent away from home is spent within the home FSA $\phi_{nt}$, and that individuals in the lowest incidence deciles spent the most time outside their home FSA (b). We aggregate $\rho_{nt}$ and $\phi_{nt}$ for each decile $g$ using the mean across corresponding FSA $n$ to yield $\rho_{gt}$ and $\phi_{gt}$.

Finally, we define the overall mobility matrix $B_{ggt}$ as

$$B_{gg't} = \rho_{gt}\left[(\phi_{gt})\delta_{gg'} + (1 - \phi_{gt})B^c_{gg't}\right] \tag{A.6}$$

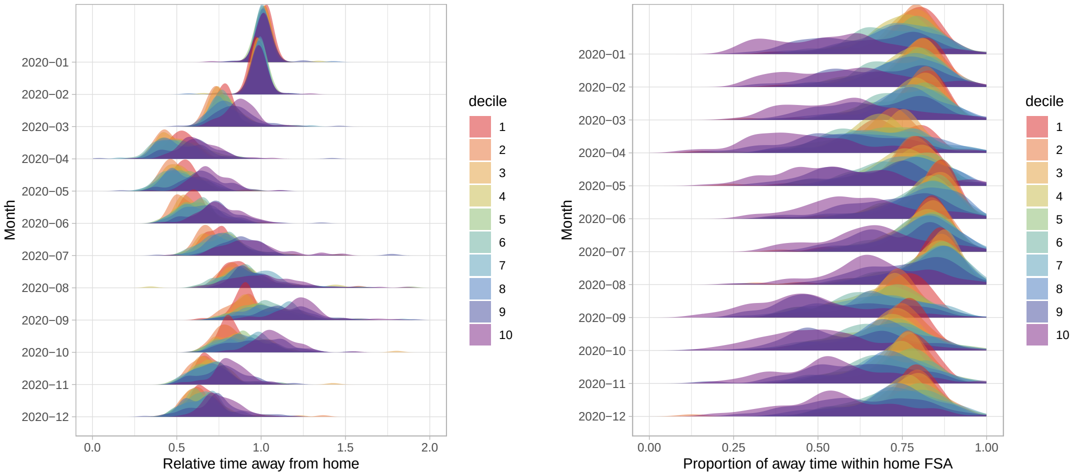


(a) Proportion of time away from home relative to the reference period ($\rho_{nt}$)

(b) Proportion of time away spent within the home FSA ($\phi_{nt}$)

Figure A.4: Measures of time away from home used to calculate the mobility matrix

Data from approximately 2% of mobile devices in Ontario, Canada; distributions show the density of ratio values across all 513 FSAs; reference period: Jan–Feb 2020.

representing a weighted average of the identity matrix $\delta_{gg'}$ and the conditional mobility matrix $B^c_{gg't}$, weighted by intra-FSA mobility ($\phi_{gt}$) and inter-FSA mobility ($1 - \phi_{gt}$), respectively, and overall proportional to $\rho_{nt}$.

##### A.3.3 Dimensionality Reduction

As written, Eq. (A.6) requires an additional observation of $B^c_{gg't}$, $\phi_{gt}$, and $\rho_{gt}$ for each month $t$. If any retrospective mobility data are missing, or for projecting $B_{gg't}$ into the future, we may be interested to know whether any of these three parameters can be extrapolated based on a subset of the original inputs.

First, we explored whether the conditional distribution of mobility destinations $B^c_{gg't}$ could be replaced by the mean across months:

$$B^c_{gg'} = \frac{1}{N_t} B^c_{gg't} \tag{A.7}$$

Figure A.5 plots the elements $(g, g')$ of $B^c_{gg't}$, showing the time trend (a) and overall distribution of values (b). Variation in $B^c_{gg't}$ between months was relatively small (coefficients of variation were: median [min (IRQ) max] = 5.4 [2.4 (4.2, 8.2) 37.9]%) suggesting that the conditional probabilities for each month could indeed be replaced by the mean as in Eq. (A.7).

Next, we explored whether the proportion of time away from home for each group and month could be replaced by the mean for each month $\rho_t$and a relative group effect $R^\rho_g$:

$$\rho_{gt} \approx \rho_t \, R^\rho_g \tag{A.8}$$

Figure A.6 plots the mean $\rho_t$ for each month, and the relative difference $R^\rho_g = \rho_{gt}/\rho_t$. Again, variation between months in $R^\rho_g$ was small (coefficients of variation: 2.8 [1.6 (2.0, 3.6) 4.2]%) suggesting that the approximation

Eq. (A.8) is reasonable. We ignore the pre-COVID-19 reference period for estimating $R_g^{\rho}$, as relative mobility $\rho_{gt} = 1$ by definition during this period.

Finally, we likewise explored whether the "intra-FSA mobility" (proportion of away time spent within the home FSA) for each group and month could be replaced by the mean for each month $\phi_t$and a relative group effect $R_g^{\phi}$:

$$\phi_{gt} \approx \phi_t \, R_g^{\phi} \tag{A.9}$$

Figure A.6 plots the mean $\phi_t$ for each month, and the relative difference $R_g^{\phi} = \phi_{gt}/\phi_t$. Again, variation between months in $R_g^{\phi}$ was small (coefficients of variation: 1.3 [0.8 (1.1, 2.5) 4.4]% ) suggesting that the approximation Eq. (A.9) is also reasonable. Since data on overall mobility $\rho_t$ may be easier to obtain or approximate than the proportion of away time spent within the home FSA $\phi_t$, we may also be interested to remove the time component of $\phi_t$ and use fixed $\phi$; in this case the approximation is worse, but still very reasonable (coefficients of variation: 7.2 [5.1 (6.5, 7.7) 11.2]% ).[13]

Thus, overall Eq.(A.6) can be re-written as:

$$B_{gg't} \approx \rho_t R_g^{\rho} \Big[ (\phi_{(t)} \, R_g^{\phi}) \delta_{gg'} + (1 - \phi_{(t)} \, R_g^{\phi}) B^c_{gg't} \Big] \tag{A.10}$$

where $\rho_t$, and possibly $\phi_t$, are the only required inputs. Using the simpler approximation with fixed $\phi$, Figure A.8 compares (a) the unapproximated mobility matrix from Eq. (A.6) with (b) the approximated mobility matrix from Eq. (A.10).

[13] It would also be helpful if $\phi_t$ could be predicted by $\rho_t$, but comparing the plots (Figures A.6a and A.7a), we see they are poorly correlated.

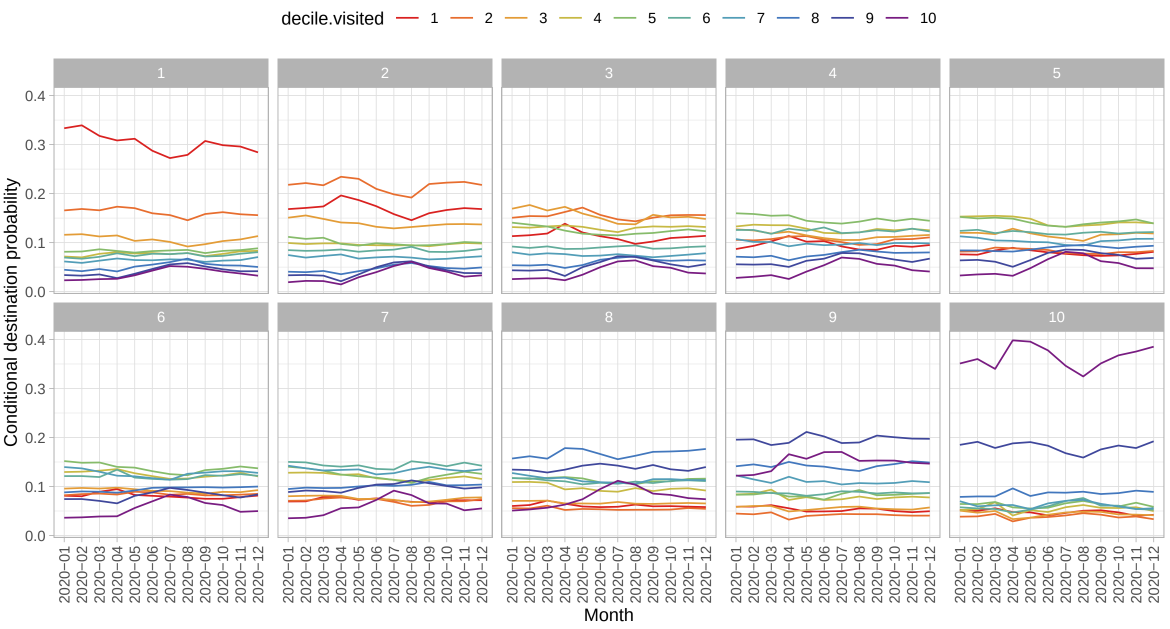


(a) Trend versus month

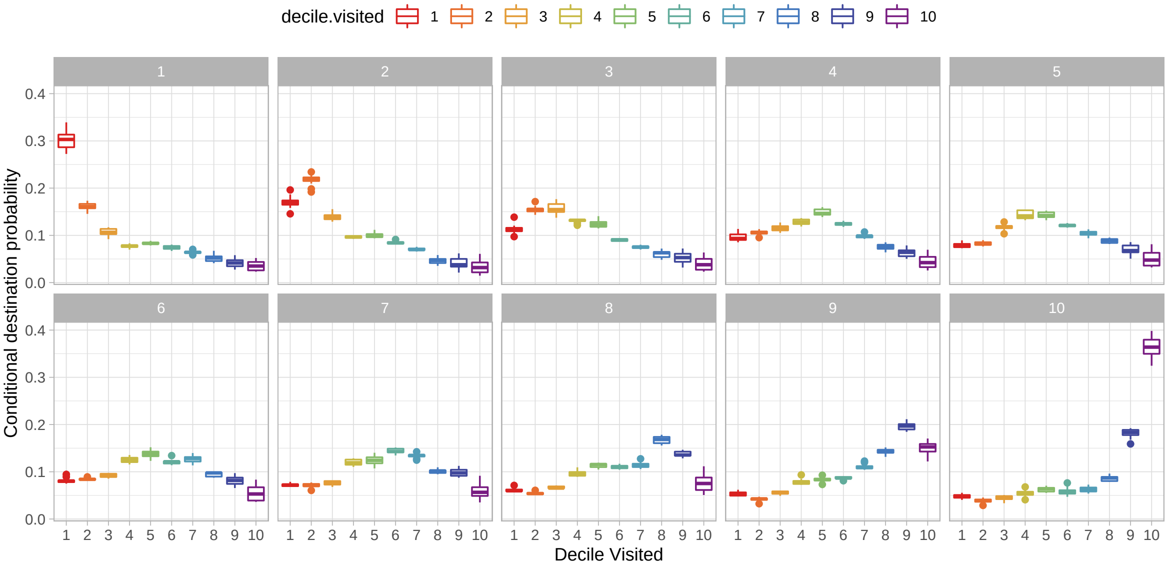


(b) Distribution of conditional probabilities

Figure A.5: Conditional probabilities of travelling from decile $g$ (panels) to decile $g'$ (colours) for Jan–Dec 2020, showing relative stability of conditional probabilities over time

Data from approximately 2% of mobile devices in Ontario, Canada.

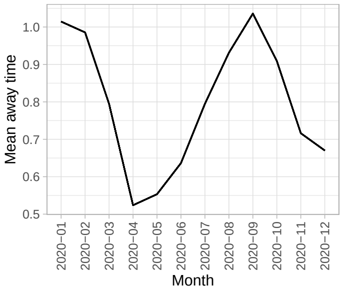


(a) Trend versus month: average mobility probability ($\rho_t$)

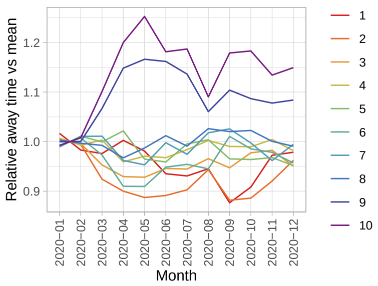


(b) Trend versus month: relative differences in mobility by decile ($\rho_{gt}/\rho_t$)

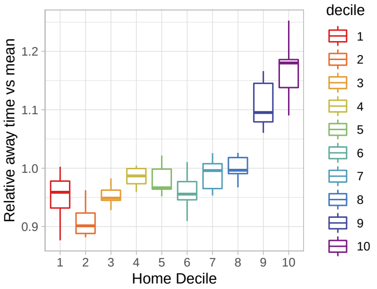


(c) Distribution of relative differences* ($\rho_{gt}/\rho_t$)

Figure A.6: Probability of being mobile for each decile $\rho_{gt}$ versus the population average, showing relative stability over time

* During March–Dec 2020 only; data from approximately 2% of mobile devices in Ontario, Canada.

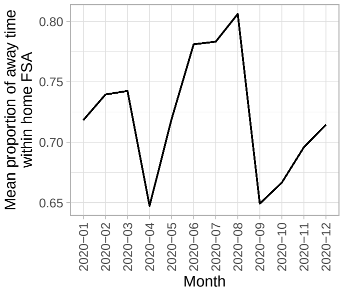


(a) Trend versus month: proportion of away time in home FSA ($\phi_t$)

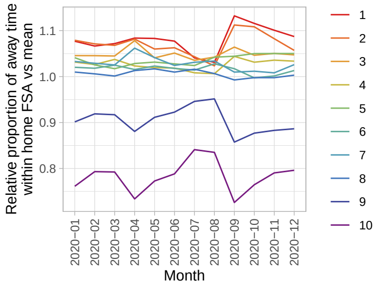


(b) Trend versus month: relative differences in proportion of away time in home FSA by decile ($\phi_{gt}/\phi_t$)

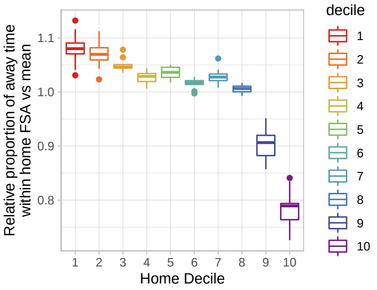


(c) Distribution of relative differences ($\phi_{gt}/\phi_t$)

Figure A.7: Conditional probability of intra-FSA mobility for each decile $\phi_{gt}$ versus the population average, showing relative stability over time

Data from approximately 2% of mobile devices in Ontario, Canada.

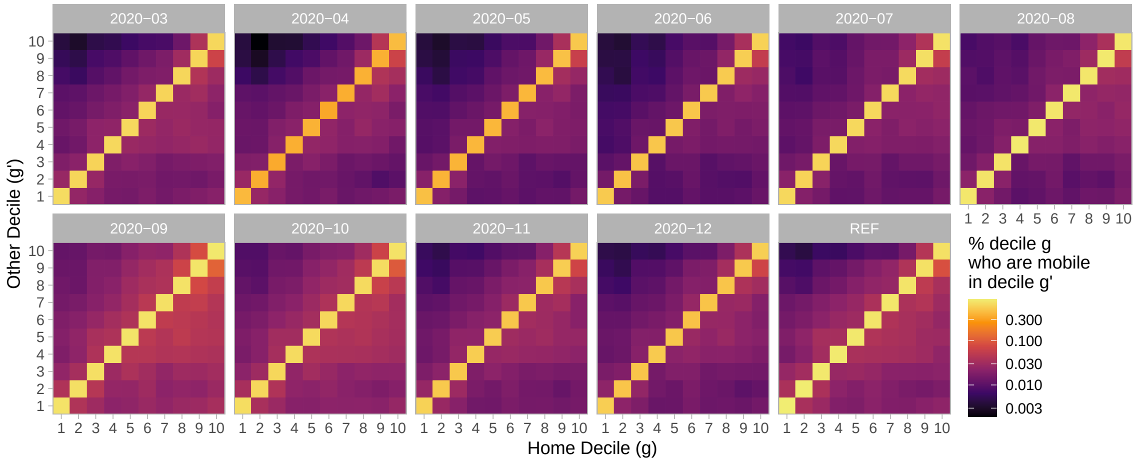


(a) Observed

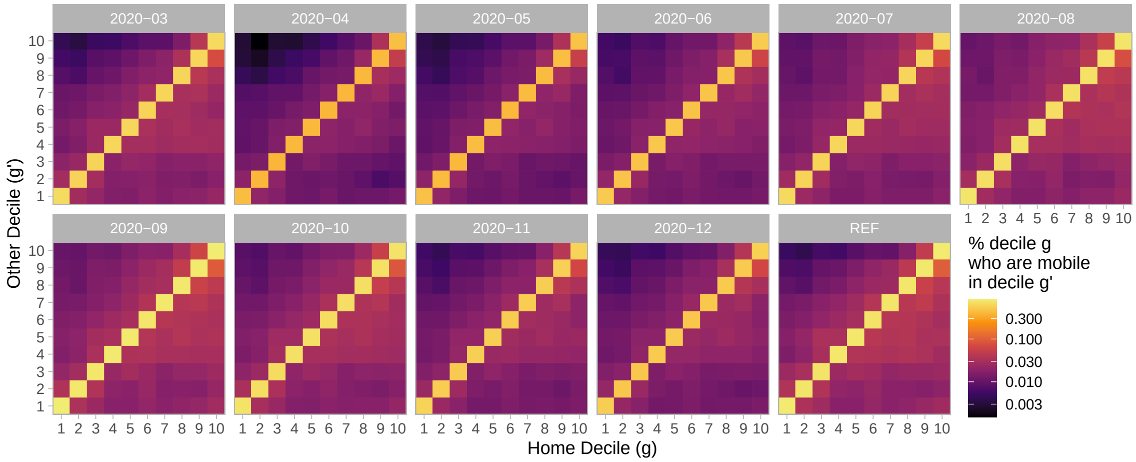


(b) Approximated

Figure A.8: Visual comparison of observed mobility matrix $B_{gg't}$, Eq. (A.6) and approximated mobility matrix, Eq. (A.10) for Mar–Dec 2020, plus REF ($t_0$) as the average of Jan–Feb 2020

Derived from mobile device geolocation data; deciles represent groupings of Ontario forward sortation areas (FSAs) by cumulative COVID-19 cases between 15 Jan 2020–28 Mar 2021; colour scale is square-root transformed to improve perception of smaller values; reference period: Jan–Feb 2020.

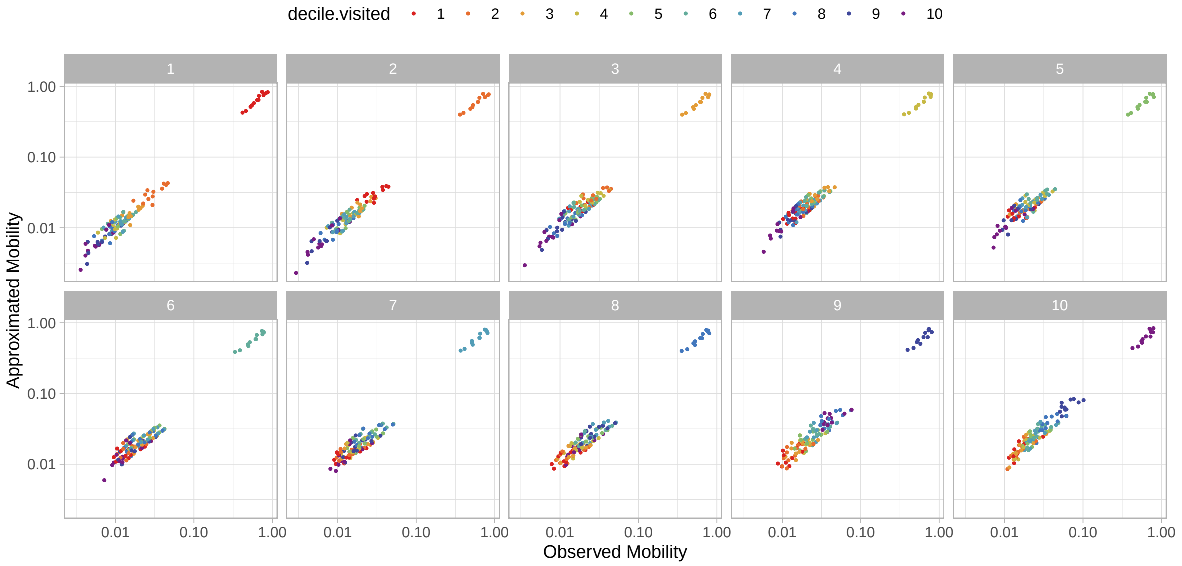


Figure A.9: Comparison of element values ($g$, panels; $g'$, colours) in the observed mobility matrix $B_{gg't}$, Eq. (A.6) and approximated mobility matrix, Eq. (A.10), showing close agreement

Derived from mobile device geolocation data; deciles represent groupings of Ontario forward sortation areas (FSAs) by cumulative COVID-19 cases between 15 Jan 2020–28 Mar 2021; x-y scales are $\log_{10}$ transformed to improve perception of smaller values

#### A.4 Additional Data for Ontario Patches

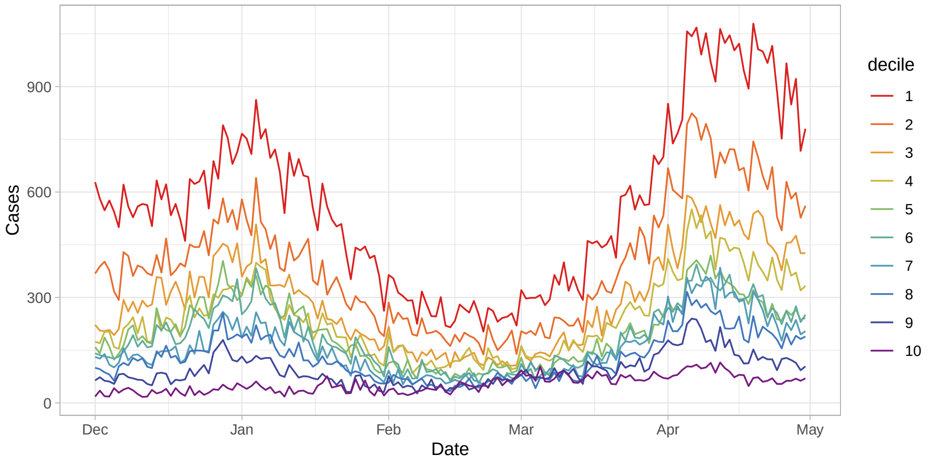


Figure A.10: Time trends in daily COVID-19 cases across the 10 deciles (patches) in Ontario

Deciles represent groupings of Ontario forward sortation areas (FSAs) by cumulative COVID-19 cases between 15 Jan 2020–28 Mar 2021.

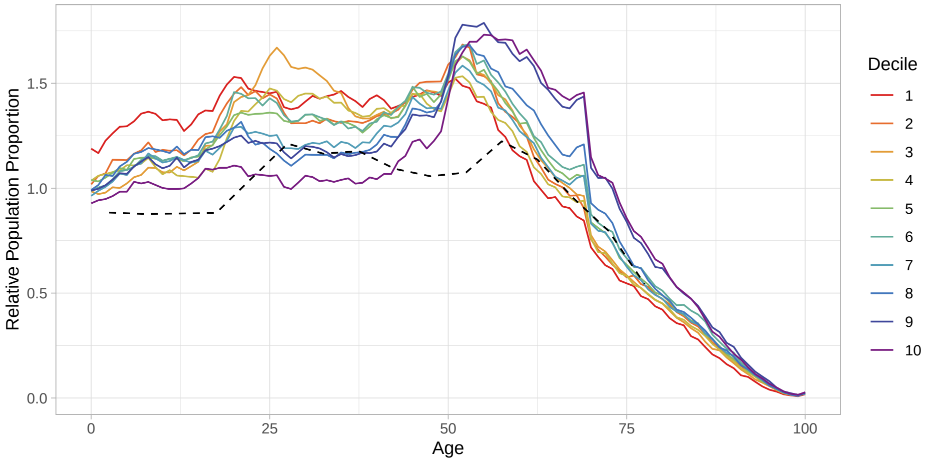


Figure A.11: Population age distributions across the 10 deciles (patches) in Ontario

Deciles represent groupings of Ontario forward sortation areas (FSAs) by cumulative COVID-19 cases between 15 Jan 2020–28 Mar 2021; solid coloured lines correspond to deciles; dashed black line corresponds to Canadian age distribution used by Prem, Cook, and Jit [2].
